# The Association of Late-Life Self-Reported Vision and Hearing Impairments with Cognitive Function: A 3-Year Longitudinal Mediation Analysis Through Social Isolation

**DOI:** 10.1101/2020.09.30.20204271

**Authors:** Jeremy B. Yorgason, Corinna Trujillo Tanner, Stephanie Richardson, Melanie S. Hill, Brian Stagg, Markus Wettstein, Joshua R. Ehrlich

## Abstract

**Background and Objectives:** Vision impairment (VI), hearing impairment (HI), and dual sensory impairment (DSI), are prevalent among older adults and have been associated with cognitive impairment in later life. Knowledge about mediating pathways that account for the association between sensory impairment and cognitive impairment is scarce. Mediators of this association could serve as possible future intervention targets. We examined longitudinal associations between sensory impairment and cognitive functioning indirectly through social isolation.

**Research Design and Methods:** Data were taken from a nationally representative panel study, The National Health and Aging Trends Study, an annual survey of Medicare beneficiaries age ≥ 65. Participants (*N* = 6,286) from Rounds 5, 6, and 7 (2015, 2016, 2017), with complete data on self-reported VI and/or HI status at baseline. Structural equation models were estimated to test longitudinal direct and indirect associations.

**Results:** Adjusting for covariates, cross-sectional results indicated that all sensory impairments (VI, HI, and DSI) were negatively associated with all cognitive functioning measures through social isolation. Longitudinally, only VI was indirectly associated with word-recall scores across 1 and 2 years through social isolation, as well as across 2 years for orientation.

**Discussion and Implications:** As social isolation is both a consequence of sensory impairment and a risk factor for cognitive impairment, it provided a starting point from which to study the process of cognitive decline among those with sensory impairments. Awareness of the association of sensory impairment with social isolation, as well as its longitudinal implications for cognitive health, may enhance our ability to intervene.

## Background and Objectives

Sensory impairments, including vision impairment (VI), hearing impairment (HI), and dual sensory impairment (DSI), are highly prevalent among older adults, and the number of individuals affected is rapidly increasing as the population ages (Swenor et al., 2013; Teutsch et al., 2016). The impact of sensory impairments on various developmental outcomes has been well documented. Self-reported vision impairment, which affects approximately 9% of adults age 65 and older in the United States (Patel et al., 2020), and self-reported hearing impairment, which affects 31% of those ages 60 to 69 and 63.1% of those age 70 and older (Goman & Lin, 2016), have a negative impact on many domains of health, functioning, and quality of life (Bainbridge & Wallhagen, 2014; Swenor et al., 2019; Teutsch et al., 2016). These challenges are compounded in the case of dual sensory impairment, which is present in up to 11% of adults age 60 and older (Correia et al., 2016; Saunders & Echt, 2007; Schneider et al., 2011; Swenor et al., 2013).

All sensory impairments increase risk for social isolation (Coyle et al., 2016; Shah et al., 2020; Shukla et al., 2020), which cross-sectional research has demonstrated to be an important mediator of the relationship between sensory impairment and cognitive impairment (Dichgans & Leys, 2017; Whitson et al., 2018). However, when considering whether the association of sensory impairments with cognitive decline occurs via social isolation, it is essential to consider longitudinal data in order to provide evidence of processes that develop over time and to establish directionality. Moreover, doing so in a sample that is representative of the population may allow for greater generalizability of findings. Therefore, the aim of this study was to investigate whether longitudinal associations between sensory and cognitive impairments were associated indirectly through social isolation in a nationally representative sample of older adults.

### The Direct Link Between Sensory Impairment and Cognitive Functioning

VI, HI, and DSI are independently associated with cognitive impairment (Lin & Albert, 2014; Lin et al., 2004; Whitson et al., 2018; Zheng et al., 2018). The relationships between sensory impairment and cognitive impairment are complex, and numerous mechanisms have been hypothesized. Sensory impairment and cognitive impairment may share common causes, such as changes of the central nervous system (Baltes & Lindenberger, 1997), vascular disease, and neurodegeneration (Dichgans, & Leys, 2017; Lupien & Lepage, 2001). Other common causes of vision impairment and cognitive impairment have been proposed, as amyloid-β, a hallmark finding in Alzheimer’s disease, has been identified in the crystalline lens and the retina of individuals with Alzheimer’s disease (Lupien & Lepage, 2001). Sensory impairments may also increase the cognitive load required for sensory processing, which may lead to poor cognitive outcomes (Pigeon et al., 2019) or result in direct alteration of brain structure, including both regional and whole-brain atrophy, due to decreased afferent sensory input (Hultsch et al., 1999; Lin et al., 2014; McEwen, 2000; Peelle et al., 2011,Whitson, 2018). Conversely, preserving sensory function may be protective of cognitive function and brain structure in both vision (Lim et al., 2020; Tamura et al., 2004; Xu et al., 2014) and hearing (Dawes et al., 2015).

### Sensory Impairment and Cognitive Functioning: The Role of Social Isolation

As noted, social isolation has been proposed as a potential mediator of the association between decreased sensory and cognitive functions (Livingston et al, 2020; Lim et al., 2020; Wahl & Heyl, 2003; Whitson et al., 2018).Vision and hearing are basic forms of interchange between individuals and their environment. When typical forms of communication are interrupted, as in the case of VI and HI, there is evidence that both social isolation (National Academies of Sciences, Engineering, and Medicine et al., 2020; Shah et al., 2020; Shukle et al, 2020) and subsequent cognitive impairment may ensue (Lim et al., 2020). Individuals with self-reported vision impairments are less likely to engage in out-of-home leisure activities (Heyl et al., 2005) or to socialize with friends or participate in social activities (Crews & Campbell, 2001). However, socially engaging activities provide cognitive stimulation, which may protect against cognitive decline (Lim et al., 2020; Lövdén et al., 2005; Whitson et al., 2018). This is consistent with the cascade hypothesis, in which cognitive decline is driven by a lack of cognitive stimulation related to sensory loss (Varadaraj, et al, 2020; Wahl & Heyl, 2003).

Both VI and HI may limit participation in activities that facilitate social engagement, such as using a telephone and driving a car (Steinman & Allen, 2012; West et al., 2002). They may limit participation in social activities, such as playing games and visiting museums (Crews & Campbell, 2001). Therefore, established literature suggests that a lack of social engagement, or *social isolation* may be a key modifiable risk factor that links sensory impairment with later cognitive impairment (Livingston, et al, 2020). Understanding these relationships may help us to identify those older adults with sensory impairments who are at increased risk of experiencing cognitive decline, as well as to design interventions directed at modifiable mechanisms.

### Current Study

In this study, we examined the longitudinal relationship between self-reported sensory impairment (i.e., HI, VI, and DSI) and cognitive impairment using a nationally representative sample from the National Health and Aging Trends Study (NHATS). We hypothesized that having one or more self-reported sensory impairments would be associated with poorer cognitive functioning cross-sectionally, and longitudinally, across 1 and 2 years and that these relationships would be associated indirectly through social isolation.

## Research Design and Methods

### Procedure and Participants

Data for the current study were taken from NHATS Rounds 5, 6, and 7 (2015-2017) (nhatsdata.org). Because these data were publicly available and were de-identified, the Institutional Review Board at Brigham Young University deemed this study exempt. NHATS is a longitudinal panel study of Medicare beneficiaries 65 years and older that began in 2011 and includes annual follow-up assessments. The sample was replenished in 2015 (Round 5), with a total sample of 8,245 persons. To decrease potential confounding and endogeneity effects in our analyses, we excluded those with probable dementia (*n* = 933), and those not living in a community setting (n=968) from the analytic sample at Round 5, thus allowing for the inference of directionality of the effect of social isolation on later cognitive impairment. Those with missing data on all predictors/covariates (*n* = 62) were also excluded, bringing the analytic sample to 6,286 individuals.

### Cognitive Measures

We employed cognitive measures collected within the NHATS study consisting of orientation, executive function, and retrieval of information (Kasper et al., 2013).

#### Orientation

NHATS interviewers asked respondents, “Without looking at a calendar or watch, please tell me today’s date,” which included month, day, year, and day of the week. Respondents were also asked to name the current president and vice president of the United States. Scores from these items measuring orientation were then summed together. Higher scores indicated greater cognitive orientation.

#### Executive Function

Executive function was assessed using a clock-drawing test. Respondents were given a sheet of paper and were instructed to draw a clock with the hands showing 11:10 (10 minutes past 11:00 o’clock). They were given two minutes to complete the activity. When finished, the drawings were scored on a scale from 0 (*not recognizable as a clock*) to 5 (*accurate depiction*). Higher scores indicated better executive function.

#### Retrieval of Information

Retrieval of information was assessed using a test of delayed word recall. Interviewers read a list of 10 nouns to the respondents before asking them to complete the clock-drawing and orientation tests. After the clock-drawing and orientation assessments were administered, respondents were asked to recall any of the nouns from the list read earlier. Higher scores indicated the recall of more words.

### Self-Reported Sensory Impairment

Three separate variables were used to assess self-reported sensory impairment: HI, VI, and DSI. A dichotomous measure of self-reported HI was constructed using four items. If the respondent reported difficulty with any of the items, they were then coded as having hearing impairment. Hearing impairment questions related to whether respondents could “hear well enough to carry on a conversation in a quiet room,” “hear well enough to carry on a conversation in a room with a radio or TV playing,” and “hear well enough to use the telephone,” and an item assessing whether participants were deaf. Response options were “Yes” (coded as 1) and “No” (coded as 0). The measure characterized people as having HI only if hearing problems were severe enough to impact their functioning (whether or not they wore a hearing aid). People with hearing aids but who did not report problems with these listed items were not coded as having a hearing impairment for this study.

Self-reported vision impairment was measured using a total of three items. If the participant reported being blind, or that they were unable to see well enough to recognize someone across the street or to read newspaper print, they were then coded as having a VI. This method was used in prior studies using NHATS data (Ehrlich et al., 2019; Frank et al., 2019; Xiang et al., 2020). Self-reported DSI was indicated if the participant reported having both HI and VI. In the current sample, 10% (*n* = 578) reported HI, 6% (*n* = 343) reported VI, and 3% (*n* = 140) reported DSI.

### Social Isolation

Social isolation was measured based on participant responses to five questions about their social network (Cudjoe et al., 2020). Participants received one point if they lived alone or if they talked to one person or fewer about “important matters” during the last year. Additionally, participants received a point each for not attending religious services and clubs/classes/organized activities and for not participating in volunteer work all during the past month. Points were summed so that higher scores indicated greater social isolation.

### Covariates

Sociodemographic characteristics were reported by respondents, including age, gender, marital status, race, education, smoking status, and chronic health conditions. Respondents reported their chronological age, gender, and marital status (not married, married). Participants indicated whether their race was “White, non-Hispanic,” “Black, non-Hispanic,” “Hispanic,” or “Other (American Indian, Native Hawaiian/Pacific Islander, Other).” Respondents were asked to indicate their highest level of attained education, with responses ranging from no schooling to master’s, professional, or doctoral degree. Education was included as an indicator of socioeconomic status (Darin-Mattsson, Fors, & Kareholt (2017). Although income is also a good indicator socioeconomic status indicator, we chose to use education as it relates to our outcome in important ways (Livingston et al., 2020), and education and income were correlated at above *r* = .50 in the current sample. Respondents reported their smoking status (nonsmoker, smoker) and whether or not they had been diagnosed with each of the following (measured as separate indicators): heart disease, hypertension, diabetes, and stroke.

### Statistical Analysis

Descriptive statistics, bivariate correlations, and mean difference tests were estimated in *STATA/SE* (StataCorp, 2019). Structural equation modeling, using *Mplus* (Muthén & Muthén, 2017), was used to test study hypotheses. Specifically, we examined associations of Round 5 sensory-impairment measures with Rounds 5 and 6 of social isolation and Rounds 5, 6, and 7 of the cognitive-functioning measures, modeling both direct and indirect associations cross-sectionally and across 1- and 2-year periods (see Figure 1). Models in Table 2 were adjusted for all covariates, including prior-wave measures of all outcome variables (see longitudinal mediation approach suggested by Lockhart, MacKinnon, & Ohlrich, 2008). VI and HI predictors were estimated together in the same models, allowing comparisons between the two. Models including DSI were estimated separately. Estimates were calculated using full-information maximum likelihood, an estimation approach that uses all available data to address missing data. Bootstrapping with 5,000 draws was used to adjust the standard errors associated with indirect effects (Hayes & Preacher, 2010).

**Figure 1.**
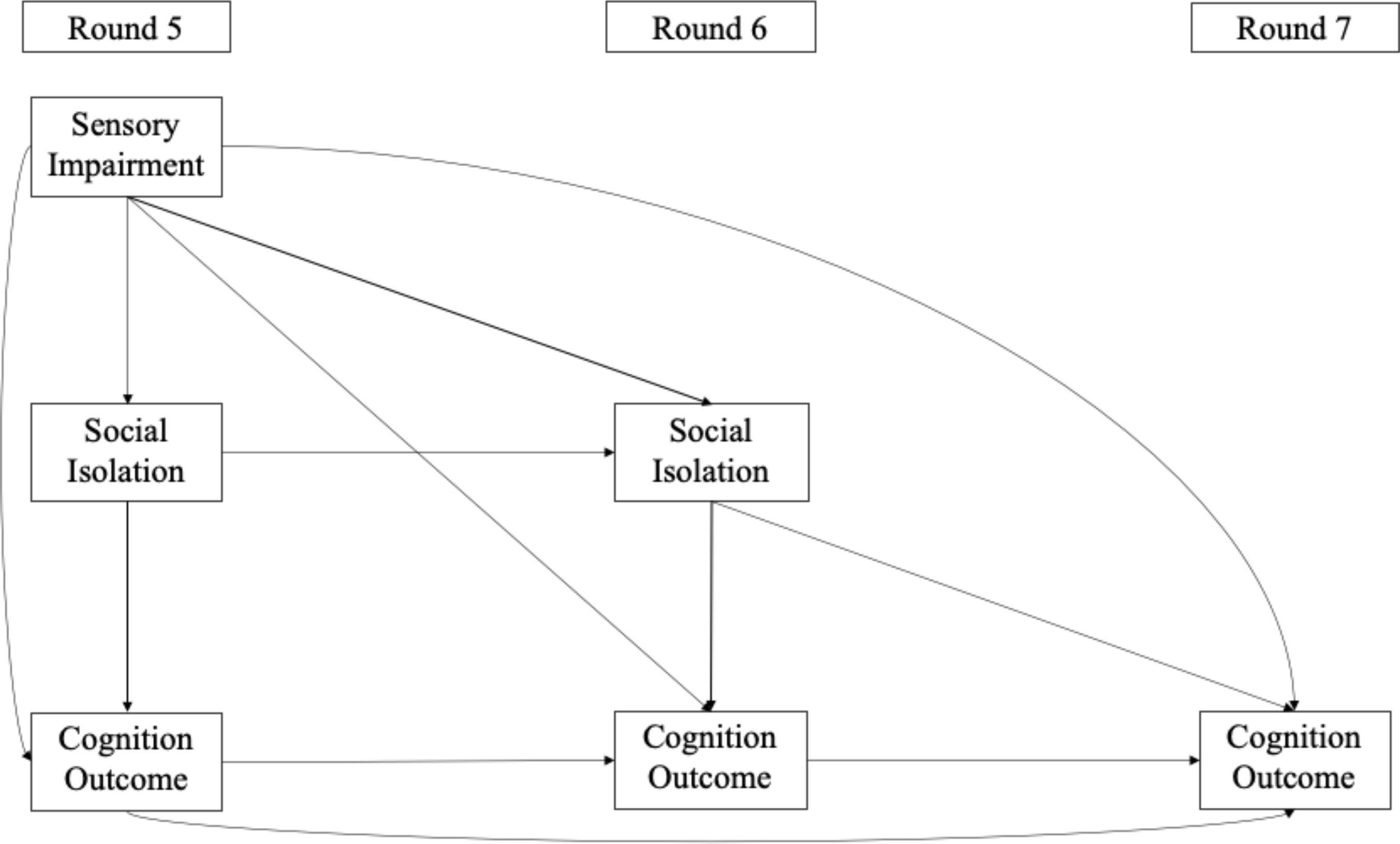
Conceptual Model Showing Direct and Indirect Pathways From Sensory Impairment to Cognitive Impairment Through Social Isolation at and Across Multiple Timepoints

**Table 1.** Descriptive Statistics and ANOVA Analyses of Main Study Variables.

| Variable | *M (SD) or n (%)* |  |  |  |  | F-Value |
| --- | --- | --- | --- | --- | --- | --- |
|  | NHATS Total Study Sample (*n* = 6,348) | No Impairment (*n* = 4,511) | Hearing Impairment (*n* = 578) | Vision Impairment (*n* = 343) | Dual Impairment (*n* = 140) |  |
| Age |  | 1.79<sup>a</sup> | 2.50<sup>b</sup> | 2.14<sup>c</sup> | 2.86<sup>d</sup> | 69.59*** |
| 0 = 65–69 | 1,013 (15.98%) | 829 (18.38%) | 68 (11.76%) | 50 (14.58%) | 18 (12.86%) |  |
| 1 = 70–74 | 1,651 (26.05%) | 1,310 (29.04%) | 109 (18.86%) | 78 (22.74%) | 17 (12.14%) |  |
| 2 = 75–79 | 1,428 (22.53%) | 1,053 (23.34%) | 111 (19.20%) | 86 (25.07%) | 22 (15.71%) |  |
| 3 = 80–84 | 1,126 (17.77%) | 753 (16.69%) | 125 (21.63%) | 55 (16.03%) | 23 (16.43%) |  |
| 4 = 85–89 | 728 (11.49%) | 410 (9.09%) | 88 (15.22%) | 52 (15.16%) | 29 (20.71%) |  |
| 5 = ≥ 90 | 392 (6.18%) | 156 (3.46%) | 77 (13.32%) | 22 (6.41%) | 31 (22.14%) |  |
| Gender |  | .40<sup>a</sup> | .52<sup>b</sup> | .33<sup>c</sup> | .37<sup>a</sup> | 13.04*** |
| 0 = Male | 3,793 (57.18%) | 2,688 (59.59%) | 279 (48.27%) | 231 (67.35%) | 88 (62.86%) |  |
| 1 = Female | 2,841 (42.82%) | 1,823 (40.41%) | 299 (51.73%) | 112 (32.65%) | 52 (37.14%) |  |
| Education |  | 4.51<sup>a</sup> | 3.99<sup>b</sup> | 3.46<sup>c</sup> | 3.15<sup>c</sup> | 44.87*** |
| 0 = No Schooling | 80 (.80%) | 13 (.29%) | 3 (.53%) | 7 (2.08%) | 2 (1.46%) |  |
| 1 = 1<sup>st</sup> - 8<sup>th</sup> grade | 1,016 (10.11%) | 291 (6.59%) | 65 (11.44%) | 61 (18.15%) | 31 (22.63%) |  |
| 2 = 9<sup>th</sup> - 12<sup>th</sup> grade | 1,307 (13.01%) | 499 (11.30%) | 76 (13.38%) | 44 (13.10%) | 28 (20.44%) |  |
| 3 = High school grad | 2,782 (27.70%) | 1,164 (26.36%) | 170 (29.93%) | 102 (30.36%) | 38 (27.74%) |  |
| 4 = Technical diploma | 740 (7.37%) | 341 (7.72%) | 36 (6.34%) | 28 (8.33%) | 8 (5.84%) |  |
| 5 = Some college | 1,398 (13.92%) | 681 (15.42%) | 81 (14.26%) | 35 (10.42%) | 9 (6.57%) |  |
| 6 = Associate degree | 469 (4.67%) | 235 (5.32%) | 22 (3.87%) | 13 (3.87%) | 2 (1.46%) |  |
| 7 = Bachelor's degree | 1,203 (11.98%) | 595 (13.47%) | 64 (11.27%) | 24 (7.14%) | 11 (8.03%) |  |
| 8 = Graduate degree | 1,050 (10.45%) | 597 (13.52%) | 51 (8.98%) | 22 (6.55%) | 8 (5.84%) |  |
| Marital Status |  | .51<sup>a</sup> | .52<sup>a</sup> | .38<sup>b</sup> | .35<sup>b</sup> | 6.44*** |
| 0 = Not married | 1,731 (49.58%) | 1,268 (49.28%) | 148 (48.21%) | 114 (62.30%) | 51 (65.38%) |  |
| 1 = Married | 1,760 (50.42%) | 1,305 (50.72%) | 159 (51.79%) | 69 (37.70%) | 27 (34.62%) |  |
| Number in Social Network | 2.18 (1.34) | 2.20<sup>a</sup> | 2.01<sup>b</sup> | 2.01<sup>a,b</sup> | 1.88<sup>b</sup> | 7.35*** |
| Number of Chronic | 2.46 (1.45) | 2.40<sup>a</sup> | 2.65<sup>b</sup> | 2.79<sup>b</sup> | 3.31<sup>c</sup> | 27.57*** |
| Illnesses |  |  |  |  |  |  |
| Depressive Symptoms | .85 (1.28) | .79<sup>a</sup> | 1.16<sup>b</sup> | 1.34<sup>b</sup> | 1.88<sup>c</sup> | 60.02*** |
| ADLs | 1.31 (1.19) | 1.12<sup>a</sup> | 1.45<sup>b</sup> | 1.57<sup>b</sup> | 2.15<sup>c</sup> | 65.62*** |
| IADLs | .79 (.93) | .72<sup>a</sup> | 1.05<sup>b</sup> | 1.12<sup>b</sup> | 1.44<sup>c</sup> | 45.19*** |
| Social Isolation R5 | 2.51 (1.27) | 2.42<sup>a</sup> | 2.72<sup>b</sup> | 2.74<sup>b</sup> | 3.12<sup>c</sup> | 26.59*** |
| Social Isolation R6 | 2.45 (1.26) | 2.36<sup>a</sup> | 2.60<sup>b</sup> | 2.78<sup>b,c</sup> | 3.09<sup>c</sup> | 23.78*** |
| Clock-Drawing R5 | 3.77 (1.07) | 3.83<sup>a</sup> | 3.69<sup>b</sup> | 3.39<sup>c</sup> | 3.16<sup>c</sup> | 33.05*** |
| Not recognizable | 36 (0.58%) | 24 (.54%) | 0 (.00%) | 6 (1.83%) | 3 (2.34%) |  |
| Severely distorted | 140 (2.24%) | 87 (1.94%) | 18 (3.16%) | 13 (3.98%) | 3 (2.34%) |  |
| Moderately distorted | 636 (10.16%) | 425 (9.50%) | 63 (11.07%) | 52 (15.90%) | 31 (24.22%) |  |
| Mildly distorted | 1,330 (21.26%) | 907 (20.26%) | 131 (23.02%) | 89 (27.22%) | 39 (30.47%) |  |
| Reasonably accurate | 2,378 (38.01%) | 1,702 (38.03%) | 220 (38.66%) | 110 (33.64%) | 38 (26.69%) |  |
| Accurate | 1,737 (27.76%) | 1,331 (29.74%) | 137 (24.08%) | 57 (17.43%) | 14 (10.94%) |  |
| Clock-Drawing R6 | 3.81 (1.08) | 3.86<sup>a</sup> | 3.71<sup>b</sup> | 3.33<sup>c</sup> | 3.33<sup>c</sup> | 29.99*** |
| Clock-Drawing R7 | 3.82 (1.09) | 3.88<sup>a</sup> | 3.72<sup>b</sup> | 3.30<sup>c</sup> | 3.21<sup>c</sup> | 31.34*** |
| Delayed Word Recall R5 | 3.61 (2.01) | 3.71<sup>a</sup> | 3.16<sup>b</sup> | 3.20<sup>b</sup> | 2.47<sup>c</sup> | 33.22*** |
| Delayed Word Recall R6 | 3.66 (2.08) | 3.79<sup>a</sup> | 3.26<sup>b</sup> | 3.00<sup>b</sup> | 2.35<sup>c</sup> | 35.54*** |
| Delayed Word Recall R7 | 3.60 (2.06) | 3.72<sup>a</sup> | 3.13<sup>b</sup> | 3.15<sup>b,c</sup> | 2.54<sup>b,c</sup> | 22.47*** |
| Orientation R5 | 6.69 (1.44) | 6.79<sup>a</sup> | 6.32<sup>b</sup> | 6.17<sup>b</sup> | 5.58<sup>c</sup> | 62.36*** |
| Orientation R6 | 6.66 (1.53) | 6.76<sup>a</sup> | 6.32<sup>b</sup> | 6.22<sup>b</sup> | 5.59<sup>c</sup> | 39.37*** |
| Orientation R7 | 6.30 (1.53) | 6.38<sup>a</sup> | 6.00<sup>b</sup> | 5.74<sup>b,c</sup> | 5.28<sup>c</sup> | 32.03*** |

*Notes.* NHATS = National Health and Aging Trends Study; ADLs = activities of daily living; IADLs = instrumental activities of daily living; R = Round. The final *N* for this table excludes those with “probable dementia” (*n* = 933) and/or who were not community-dwelling (*n* = 968) but includes those with missing on covariates (*n* = 62). R5 was in 2015, R6 was in 2016, and R7 was in 2017. Demographic characteristics were measured at R5 of NHATS. Superscripts a-d represent significant group differences. \*\*\**p* < .001.

**Table 2.** Standardized Regression Coefficients From Structural Equation Model of Self-Reported **Vision and Hearing** Impairment Predicting Cognitive Functioning Indirectly Through Social Isolation

| Predictor Variable | Model 1<br>Word Recall<br>$\beta$ (SE) | Model 2<br>Orientation<br>$\beta$ (SE) | Model 3<br>Clock Drawing<br>$\beta$ (SE) |
| --- | --- | --- | --- |
| VI → R5 CI | -.013 (.01) | -.054*** (.01) | -.083*** (.01) |
| VI → R6 CI | -.031** (.01) | -.007 (.01) | -.062*** (.01) |
| VI → R7 CI | .015 (.01) | -.014 (.01) | -.040** (.02) |
| HI → R5 CI | -.036** (.01) | -.058*** (.01) | -.003 (.01) |
| HI → R6 CI | -.006 (.01) | -.012 (.01) | .008 (.01) |
| HI → R7 CI | -.014 (.01) | .005 (.01) | .013 (.02) |
| Social Isolation → R5 CI | -.091*** (.01) | -.068*** (.01) | -.055*** (.01) |
| Social Isolation → R6 CI | -.081*** (.01) | -.029* (.01) | -.032* (.01) |
| Social Isolation → R7 CI | -.023* (.01) | -.037** (.01) | -.023± (.01) |
| VI → R5 Social Isolation | .043*** (.01) | .033** (.01) | .032** (.01) |
| VI → R6 Social Isolation | .030** (.01) | .028** (.01) | .028** (.01) |
| HI → R5 Social Isolation | .048*** (.01) | .050*** (.01) | .056*** (.01) |
| HI → R6 Social Isolation | -.002 (.01) | .000 (.01) | .001 (.01) |
| VI → R5 Social Isolation → R5 CI | -.004** (.00) | -.002* (.00) | -.002* (.00) |
| VI → R6 Social Isolation → R6 CI | -.002** (.00) | -.001± (.00) | -.001± (.00) |
| VI → R6 Social Isolation → R7 CI | -.001* (.00) | -.001* (.00) | -.001 (.00) |
| HI → R5 Social Isolation → R5 CI | -.004** (.00) | -.003** (.00) | -.003** (.00) |
| HI → R6 Social Isolation → R6 CI | .000 (.00) | .000 (.00) | .000 (.00) |
| HI → R6 Social Isolation → R7 CI | .000 (.00) | .000 (.00) | .000 (.00) |
| Sample Size (N) | 6,286 | 6,286 | 6,286 |
| Chi-Square Model Fit | 12,093.09*** | 12,192.33*** | 8,654.02*** |
| RMSEA | .02 | .02 | .02 |
| CFI | 1.00 | 1.00 | 1.00 |
| R<sup>2</sup> of R7 CI | .47*** | .45*** | .29*** |

## Results

### Characteristics of Study Sample

Descriptive statistics of our sample, including mean-difference tests, are found in Table 1, with bivariate correlations between our main study variables found in Supplemental Table 1. At Round 5 of NHATS, participants ranged in age from 65 to 89, with the largest proportion in the 65 to 69 age range (42.03%). The majority were male (57.18%) and married (50.42%). Those with DSI were older and reported higher social-isolation scores than those with no impairment or those with only HI or VI. In contrast, those with no impairment or HI did not differ from each other in cognitive functioning but had higher cognitive-functioning scores than those with VI or DSI.

### Vision and Hearing Impairment

As seen in Table 2, self-reported VI at Round 5 was significantly negatively associated with delayed word-recall scores 1 year later (Round 6: β = –.03, *p* < .01), with concurrent orientation scores (Round 5: β = –.05, *p* < .001), and with all three waves of clock-drawing scores (Round 5: β = –.08, *p* < .001; Round 6: β = –.06, *p* < .001; Round 7: β = –.04, *p* < .01). HI at Round 5 was negatively associated with concurrent word-recall (Round 5: β = –.04, *p* < .01) and orientation scores (Round 5: β = –.06, *p* < .001). Higher social-isolation scores at Round 5 were significantly associated with lower scores on nearly all cognitive indicators at all waves. Also, VI and HI were consistently and significantly associated with higher concurrent social-isolation scores. Round 5 VI was associated with significantly higher social-isolation scores across 1 year. In contrast, HI at Round 5 was not associated with Round 6 social-isolation scores.

Results from the analysis of indirect associations (shown as *a*b)* indicated significant relationships between sensory impairments and cognitive outcomes through social isolation. Prior to adjusting for covariates, all indirect associations at all waves between VI and cognitive-functioning were statistically different from zero, and indirect associations between HI and cognitive functioning were only significant cross-sectionally (not shown). As depicted in Table 2 and in Figures 2 and 3, after adjusting for covariates, self-reported VI at Round 5 was significantly associated with all three concurrent (Round 5) cognitive-functioning measures indirectly through social isolation (word recall: *a*b* = –.004, *p* < .01; orientation: *a*b* = –.002, *p* < .05; clock drawing: *a*b* = –.002, *p* < .05). Similarly, self-reported HI at Round 5 was indirectly associated with all three concurrent (Round 5) cognitive-functioning measures through social isolation (word recall: *a*b* = –.004, *p* < .01; orientation: *a*b* = –.003, *p* < .01; clock drawing: *a*b* = –.003, *p* < .01). Longitudinal results indicated that self-reported VI at Round 5 was significantly associated with measures of word recall (*a*b* = –.002, *p* < .01) 1 year later through Round 6 social isolation, and with measures of word recall (*a*b* = –.001, *p* < .01) and orientation (*a*b* = –.001, *p* < .01) 2 years later through Round 6 social isolation. Relationship effect sizes were interpreted as follows: small = .01 to .09; medium = .09 to .24; large = ≥ .25 (Kenny, 2012).

**Figure 2.**
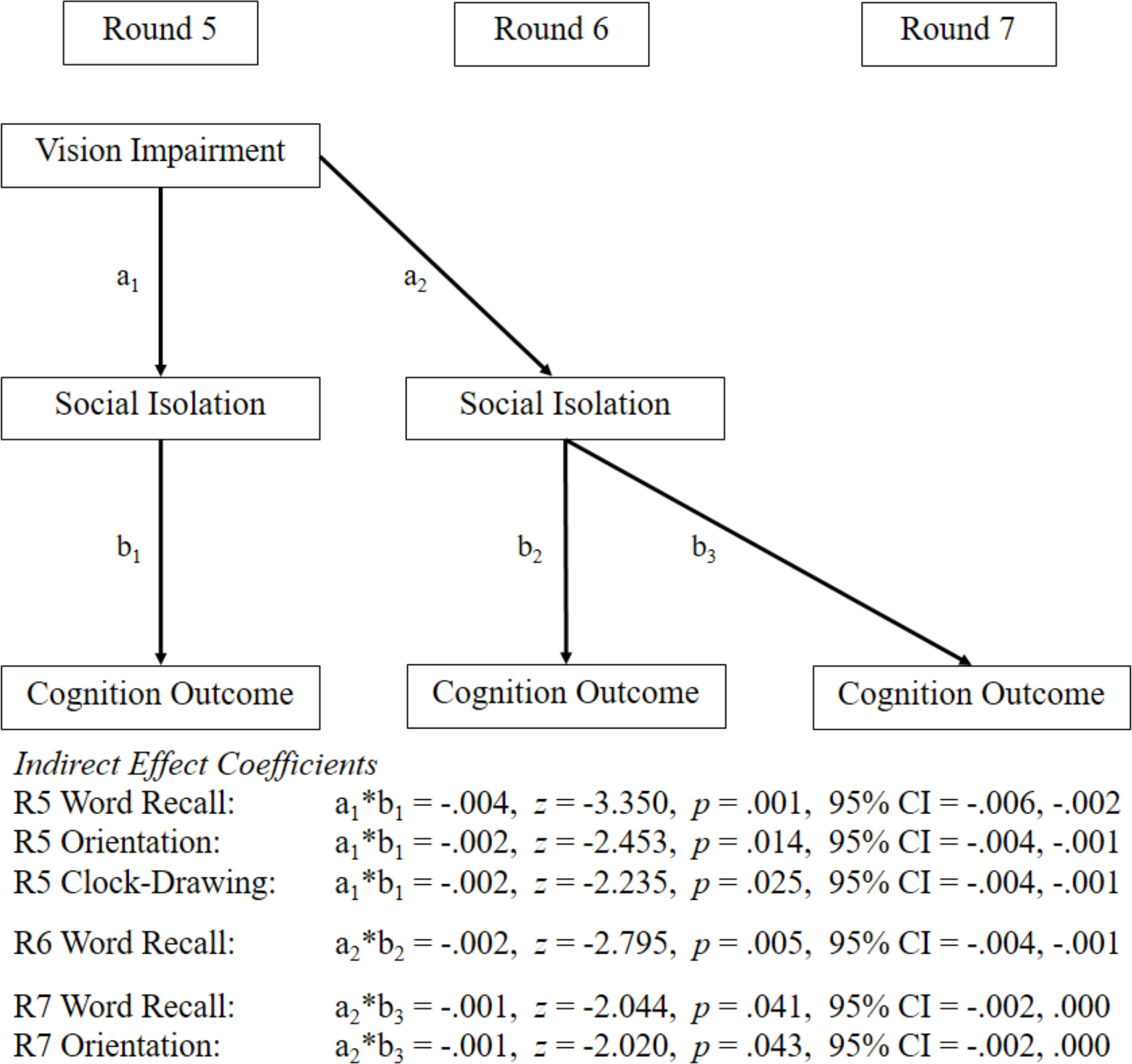
Results From Tests of Indirect Effects of Vision Impairment on Cognitive Outcomes Through Social Isolation *Notes*. R = Round of NHATS study; CI = confidence interval.

**Figure 3.**
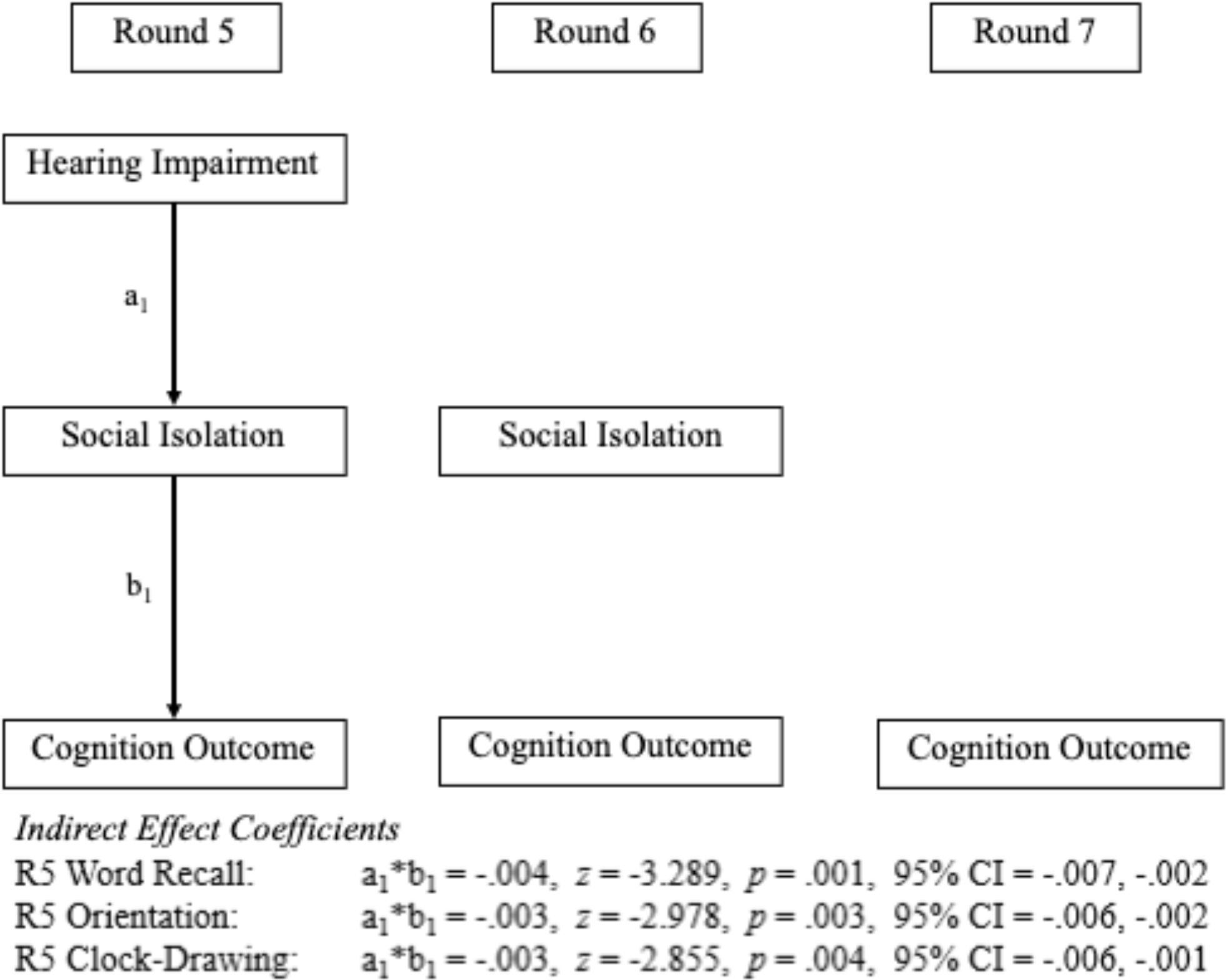
Results From Tests of Indirect Effects of Hearing Impairment on Cognitive Outcomes Through Social Isolation *Notes*. R = Round of NHATS study; CI = confidence interval.

**Figure 4.**
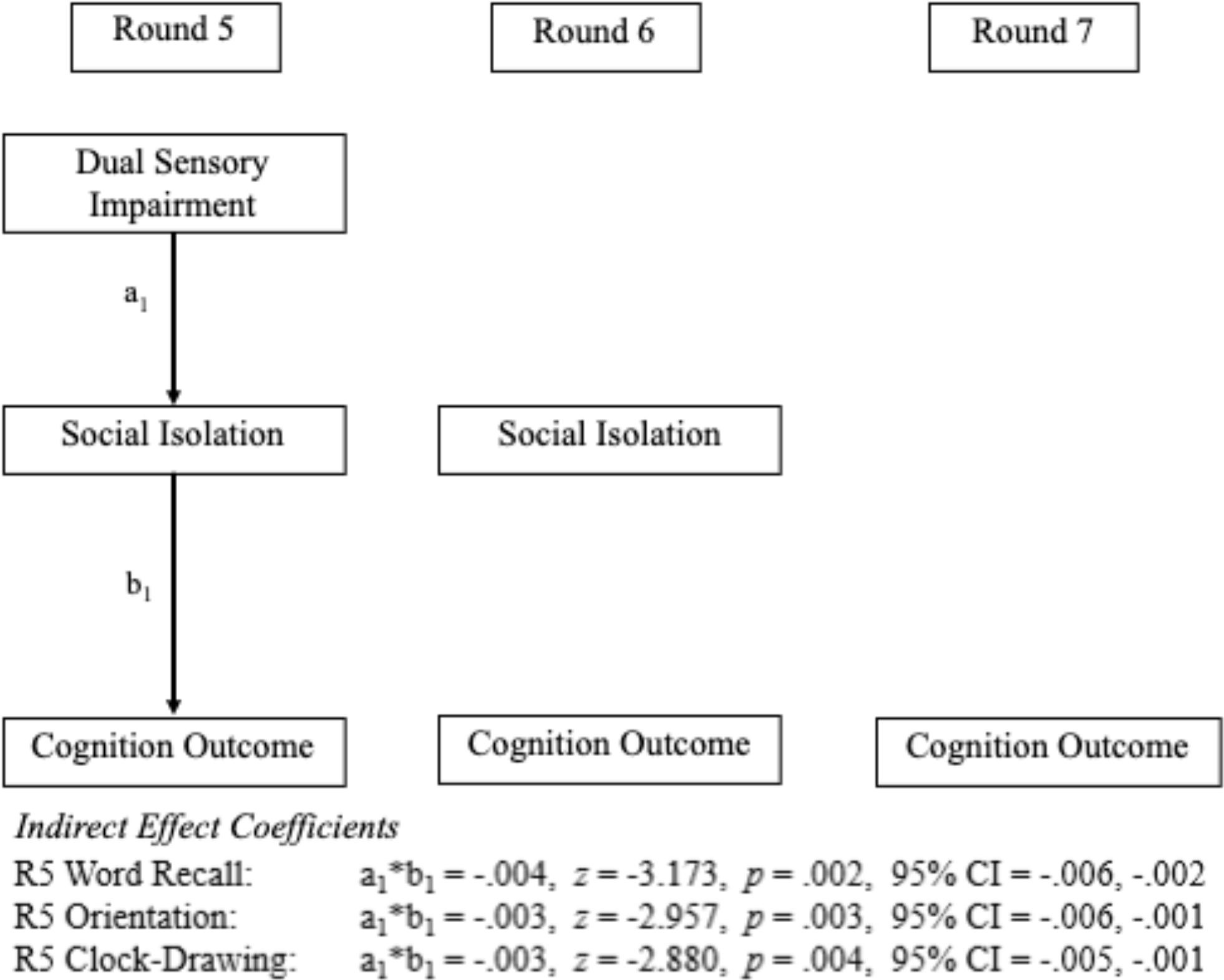
Results From Tests of Indirect Effects of Dual Sensory Impairment on Cognitive Outcomes Through Social Isolation *Notes*. R = Round of NHATS study; CI = confidence interval.

### Dual Sensory Impairment

As seen in Table 3, DSI at Round 5 was significantly related to concurrent lower word-recall (Round 5: β = –.028, *p* < .05), orientation (Round 5: β = –.055, *p* < .001), and clock-drawing scores (Round 5: β = –.048, *p* < .01). Similar to the HI/VI models, social-isolation scores at Round 5 were significantly associated with all cognitive-impairment scores at all waves in the DSI models. DSI was consistently and significantly associated with higher concurrent social-isolation scores. Regarding indirect associations, DSI at Round 5 was significantly associated with all three concurrent cognitive measures through social isolation (word recall: *a*b* = –.004, *p* < .01; orientation: *a*b* = –.003, *p* < .01; clock drawing: *a*b* = –.003, *p* < .01). DSI was not associated with cognitive functioning longitudinally. As in the HI/VI models, DSI and other predictors explained between 29% and 47% of the variance in cognitive scores at Round 7.

**Table 3.** Standardized Regression Coefficients From Structural Equation Models of Self-Reported Dual Sensory Impairment Predicting Cognitive Functioning Indirectly Through Social Isolation

| Predictor Variable | Model 1<br>Word Recall<br>$\beta$ (SE) | Model 2<br>Orientation<br>$\beta$ (SE) | Model 3<br>Clock Drawing<br>$\beta$ (SE) |
| --- | --- | --- | --- |
| DSI → R5 CI | -.028* (.01) | -.055*** (.01) | -.048** (.02) |
| DSI → R6 CI | -.018 (.01) | -.013 (.01) | -.011 (.02) |
| DSI → R7 CI | .005 (.01) | .003 (.02) | -.010 (.02) |
| Social Isolation → R5 CI | -.082*** (.01) | -.071*** (.01) | -.063*** (.01) |
| Social Isolation → R6 CI | -.086*** (.01) | -.029* (.01) | -.040** (.01) |
| Social Isolation → R7 CI | -.033** (.01) | -.038** (.01) | -.030* (.01) |
| DSI → R5 Social Isolation | .046*** (.01) | .046*** (.01) | .046*** (.01) |
| DSI → R6 Social Isolation | .020± (.01) | .020 (.01) | .020± (.01) |
| DSI → R5 Social Isolation → R5 CI | -.004** (.00) | -.003** (.00) | -.003** (.00) |
| DSI → R6 Social Isolation → R6 CI | -.002± (.00) | -.001 (.00) | -.001 (.00) |
| DSI → R6 Social Isolation → R7 CI | -.001 (.00) | -.001 (.00) | -.001 (.00) |
| Sample Size (N) | 6286 | 6286 | 6286 |
| Chi-Square | 12,313.84*** | 12,140.92*** | 8,969.64*** |
| RMSEA | .02 | .02 | .03 |
| CFI | 1.00 | 1.00 | 1.00 |
| R<sup>2</sup> of R7 CI | .47*** | .45*** | .29*** |

*Note.* DSI = dual sensory impairment, CI = cognitive impairment; RMSEA = root mean squared error of approximation, CFI = Comparative Fit Index; R = Round. R5 was in 2015, R6 was in 2016, and R7 was in 2017. Arrows denote a directional regression; multiple arrows denote an indirect relationship.

## Discussion and Implications

Using data from NHATS, we conducted a series of longitudinal analyses to test the hypothesis that the association between self-reported sensory impairments and multiple measures of cognitive impairment are mediated by social isolation. Findings partially supported this hypothesis, as all cross-sectional and select longitudinal indirect associations between VI, HI, DSI, and cognitive impairment operated through social isolation. One- and 2-year longitudinal findings indicated significant indirect associations of VI and word recall that were mediated through social isolation. Two-year longitudinal associations were also found for VI and orientation scores through social isolation. Standardized coefficients in Tables 2 and 3 suggest that sensory impairments have a small (all a*b coefficients < .01) yet significant association with cognitive functioning through social isolation. Of note, results from these models account for between 29% and 47% of the variance in cognitive-functioning scores at Round 7, suggesting that the current models provide a robust view of cognitive functioning in context of important related factors. When considering the role that social isolation plays in these associations, it is helpful to examine the decomposition of direct, indirect, and total effects. As seen in Supplemental Table 2, the proportion of indirect to total effects suggests that social isolation accounts for between 2% and 50% of the total effects of sensory impairment and cognitive functioning.

### Vision Impairment and Cognitive Impairment Through Social Isolation

Evidence in support of a longitudinal mediation effect in this study was found for VI. These findings are in line with previous research that has shown the direct associations of self-reported VI with cognitive decline among older adults cross-sectionally and longitudinally (Davies-Kershaw et al., 2018; Lin et al., 2004; Maharani et al., 2018). Similar findings have also been reported for objectively measured VI in prior studies (Lee et al., 2020; Nael et al., 2019; Zheng et al., 2018).

The current study contributes to a limited but growing literature that supports the notion that visual and cognitive impairments are associated both concurrently and over time through social interactions. Longitudinally, VI was associated with cognitive functioning two years later, through social isolation. Although approximately 50% of the variance in cognition measures was left unexplained by our models, and could be accounted for by other known determinants of cognitive ability including genetics, cognitive reserve and lifestyle factors, additional mechanistic hypotheses and mediators could be explored to further account for the longitudinal association between self-reported VI and cognitive function.

Few prior studies have sought to test the hypothesized pathway between sensory impairments and cognitive functioning through social isolation. A cross-sectional study in Canada reported only weak mediating effects by social isolation between objectively measured sensory and cognitive impairments (Hämäläinen et al., 2019). Hämäläinen and colleagues (2019) did find that social factors were most important for cognitive abilities, for females and older individuals, the latter of which was not surprising, as the sample ranged in age from 45 to 85 years. Notably, that study analyzed mediation by combining results from multiple regression equations rather than using a framework such as structural equation modeling, which estimates all equations simultaneously, provides overall model fit information, and is better suited to estimating variance in the presence of missing data (see Gunzler et al., 2013). The current study extends those findings and provides longitudinal evidence supporting these associations. Although we accounted for age, gender and other covariates, we did not explore how these characteristics might moderate direct and indirect associations. Future research is needed to better understand the specific groups for whom these associations may be most important. For instance, associations between sensory impairments and cognition are found to be stronger when individuals have higher neuroticism scores (Gaynes et al., 2013; Wettstein et al., 2016).

### Hearing Impairment and Cognitive Impairment Through Social Isolation

As hypothesized, and supported by prior research (Lin & Albert, 2014), HI was associated with higher levels of cognitive impairment through social isolation. Established literature supports similar cognitive implications for VI (Lin et al 2004; Reyes-Ortiz, 2005). HI may impede social interactions, which may lead to decreased cognitive functioning, as evident in previous research (Goman & Lin, 2016; Shukla et al., 2020) However, in the current study, HI had no longitudinal relationship with cognitive impairment through social isolation. One possible explanation is that assistive listening devices, such as hearing aids, are effective and accessible to many, allowing for adjustment to hearing impairment. Another explanation is that of under reporting of HI in the current sample. Some have found that older adults tend to *overestimate* their hearing ability (Bainbridge & Wallhagen, 2014), so that individuals in the current sample with no self-reported hearing deficits may consist of individuals with and without objective hearing loss. Another possibility is that HI acts on cognition through other indirect pathways (e.g., Wayne & Johnsrude, 2015). That is, HI, VI, and cognitive impairment may share a common etiology. Alternatively, HI and VI may lead to increased cognitive load, which may compromise cognitive functioning. Future research could combine multiple hypothesized mediators and causal factors so that their relative contributions can be quantified and compared (see Pronk et al., 2019).

### Dual Sensory Impairment and Cognitive Impairment Through Social Isolation

In partial support of our hypothesis, DSI was negatively associated with cognitive impairment through social isolation cross-sectionally. Based on our findings indicating that VI and HI are independently associated with cognitive impairment, it was not surprising that DSI was also associated with cognitive impairment. This finding is in line with prior research suggesting that DSI may be a risk factor for cognitive decline and dementia in later life (Hwang et al., 2020; Whitson et al., 2018).

Findings from the current study raise important questions about why sensory impairments have such robust concurrent associations with cognitive impairment through social isolation, yet after adjusting for likely confounders, only VI was associated longitudinally through social isolation. It appears that sensory impairments have an impact on concurrent social interactions and cognitive performance. However, it is possible that older adults with HI make adaptations to overcome social isolation, which may explain why these associations did not persist over time. Longitudinal associations may have been undetected among those with DSI because of the smaller, potentially under powered sample size. Moreover, some individuals may (partially) recover from sensory impairments, e.g. by getting hearing aids. It might be the degradation of sensory functioning over time, rather than sensory status assessed at one point I time, that drives changes in social isolation and cognitive abilities. Research examining nuances of how social networks, social connectedness, and loneliness are impacted by different sensory impairments over a long period of time is needed to better understand the potential mediating role of these psychosocial constructs on cognitive outcomes.

### Strengths

Although prior studies have reported an association between sensory and cognitive impairments, the pathways that account for this association, particularly when considered longitudinally, have not been rigorously tested. A key strength of this study is that it investigated the associations between sensory impairments and cognitive abilities as well as the potentially mediating role of social isolation longitudinally. Future research should consider other hypothesized mediators, the effect of changes in exposures, mediators, and of sensory abilities over time, and differences based on whether sensory function is assessed objectively or by self-report. In the current study, a key strength was its external validity, as data from NHATS are representative of the entire Medicare-eligible U.S. population age 65 years and older in the, which allows findings to be generalized to a broad population.

### Limitations

The current study had several limitations. First, data on sensory impairment and social isolation were based on self-reports, which may be subject to recall and social-desirability biases. Also, self-reported measures of sensory impairment may in fact represent distinct latent constructs from objective measures of sensory status (e.g., visual acuity, pure-tone audiometry). However, both types of measures may be important in assessing functional status and disability outcomes related to sensory health (Klein et al., 1999) and there is substantial overlap between self-reported and objectively measured sensory function (Cavazzana et al., 2018; Coyle et al., 2016; Pinto et al., 2014).Yet, in future work it will be important to compare associations between cognitive function and these two types of measures. Furthermore, the mediating pathways between cognitive decline and self-reported and objective sensory measures may vary. Prior research suggests that self-reported measures may be more strongly associated with certain indicators of disability (Donoghue et al., 2014; Pinto, et al, 2014) and quality of life (Bookwala, J., & Lawson, B. (2011); Gopinath et al, 2012), including social isolation (Coyle et al., 2016), whereas objective measures may be more strongly correlated with some biological processes (Warrian et al., 2010). Finally, there may have been selective dropout, wherein the least healthy participants were less likely to continue study participation and contribute complete data. This could have led to an underestimation of the true rates of cognitive decline (and a further factor that might have contributed to the underestimation are practice effects; see Salthouse, 2015) and biased results toward the null hypothesis, as survival and general health may be negatively associated with sensory impairment, social isolation, and cognitive function.

Although our results yielded significant findings, stronger associations between sensory impairment and cognitive impairment, as well as stronger mediating effects of social isolation, might have been found for other measures of cognitive outcomes. For instance, information processing speed is a key marker of cognitive functioning and of cognitive aging (Finkel et al, 2007) and has been found to be predicted by vision impairment (Chen et al., 2017). Future research may benefit from examining other cognitive outcomes in relation to VI, HI, and DSI.

### Implications

Social relationships are essential to well-being and are central to the maintenance of health. Individuals who are socially isolated are at increased risk, not only for cognitive decline (Dichgans & Leys, 2017; Whitson et al., 2018), but also for the development of other negative health outcomes, including cardiovascular disease (Barth et al., 2010) and even increased rates of mortality (National Academies of Sciences, Engineering, and Medicine et al., 2020). Social isolation is known to be more common in older adults with sensory impairment(s) (Shah et al., 2020; Shukla et al., 2020), and this challenge is now exacerbated, as many are living with restrictions related to the SARS-CoV-2 global pandemic.

Although the positive consequences of targeting social isolation as a modifiable risk factor extend beyond the scope of this study, a recent report by the *Lancet* Commission estimated that the prevalence of dementia could be decreased by 4% if social isolation were eliminated as a risk factor (Livingston et al., 2020). Globally, based on current projections, this could equate to more than four million averted cases by 2050. The findings of the current study underscore the importance of efforts to mitigate social isolation among older adults with sensory impairments. Interventions that target social isolation may have broad positive implications.

Successful aging is multidimensional and complex (Wahl, et al, 2013) and may be more challenging for older adults with sensory impairments (Swenor et al., 2019). Early diagnosis and attentive medical treatment of conditions which lead to sensory loss should be prioritized, to mitigate loss and restore function, to the greatest degree possible. Still, there is a need for interventions that go beyond medical treatments and that take a “psycho-ophthalmological perspective”, e.g. by helping visually impaired individuals to reduce stress, adjust to sensory loss, and remain socially connected (Sabel et al, 2018; Heyl & Wahl, 2014; Wahl, 2013). The positive impact of rehabilitation cannot be overstated as it focuses the use of technology and alternate techniques to ameliorate the challenges posed by sensory loss and may include low-vision rehabilitation, orientation, and mobility training for those with VI, and access to hearing rehabilitation and hearing aids for those with HI.

## Conclusion

In conclusion, this study provides important evidence for mediation by social isolation on the association between self-reported sensory impairments, especially VI, and cognitive impairment over time. The study supports the existence of longitudinal associations between both self-reported sensory impairment and social isolation with cognitive impairment in a large, nationally representative sample. Findings indicate the need for future research to test additional hypothesized mediating pathways, strategies to reduce social isolation in older adults with sensory impairment, particularly vision loss, and, ultimately, to conduct trials to determine whether cognitive decline may be mitigated through interventions to optimize sensory and social function.

## Data Availability

NHATS data are publicly available.

https://www.nhats.org/

---

**Supplemental Table 1.** Correlations Between Study Variables

| Variable | 1 | 2 | 3 | 4 | 5 | 6 | 7 | 8 | 9 | 10 | 11 | 12 | 13 | 14 |
| --- | --- | --- | --- | --- | --- | --- | --- | --- | --- | --- | --- | --- | --- | --- |
| 1. VI R5 | .14 | -- |  |  |  |  |  |  |  |  |  |  |  |  |
| 2. HI R5 | .12 | .15 | -- |  |  |  |  |  |  |  |  |  |  |  |
| 3. DSI R5 | .12 | .52 | .42 | -- |  |  |  |  |  |  |  |  |  |  |
| 4. Social Isolation R5 | .17 | .09 | .09 | .08 | -- |  |  |  |  |  |  |  |  |  |
| 5. Social Isolation R6 | .16 | .10 | .08 | .08 | .72 | -- |  |  |  |  |  |  |  |  |
| 6. Clock-Drawing R5 | -.13 | -.13 | -.06 | -.09 | -.14 | -.15 | -- |  |  |  |  |  |  |  |
| 7. Clock-Drawing R6 | -.12 | -.13 | -.05 | -.06 | -.11 | -.15 | .42 | -- |  |  |  |  |  |  |
| 8. Clock-Drawing R7 | -.14 | -.14 | -.06 | -.08 | -.13 | -.16 | .39 | .43 | -- |  |  |  |  |  |
| 9. Delayed Recall R5 | -.14 | -.09 | -.11 | -.09 | -.20 | -.19 | .27 | .28 | .30 | -- |  |  |  |  |
| 10. Delayed Recall R6 | -.15 | -.12 | -.10 | -.09 | -.20 | -.23 | .27 | .31 | .32 | .57 | -- |  |  |  |
| 11. Delayed Recall R7 | -.14 | -.08 | -.10 | -.07 | -.18 | -.19 | .25 | .27 | .34 | .56 | .61 | -- |  |  |
| 12. Orientation R5 | -.16 | -.14 | -.13 | -.12 | -.18 | -.18 | .23 | .24 | .26 | .37 | .38 | .35 | -- |  |
| 13. Orientation R6 | -.16 | -.12 | -.11 | -.10 | -.17 | -.18 | .23 | .27 | .27 | .39 | .42 | .40 | .65 | -- |
| 14. Orientation R7 | -.18 | -.13 | -.09 | -.09 | -.18 | -.18 | .26 | .28 | .33 | .40 | .42 | .45 | .54 | .61 |

*Note.* Symp. = symptoms; R = Round; VI = vision impairment; HI = hearing impairment; DSI = dual sensory impairment.
All coefficients are significant at $p < .001$ .

**Supplemental Table 2.**
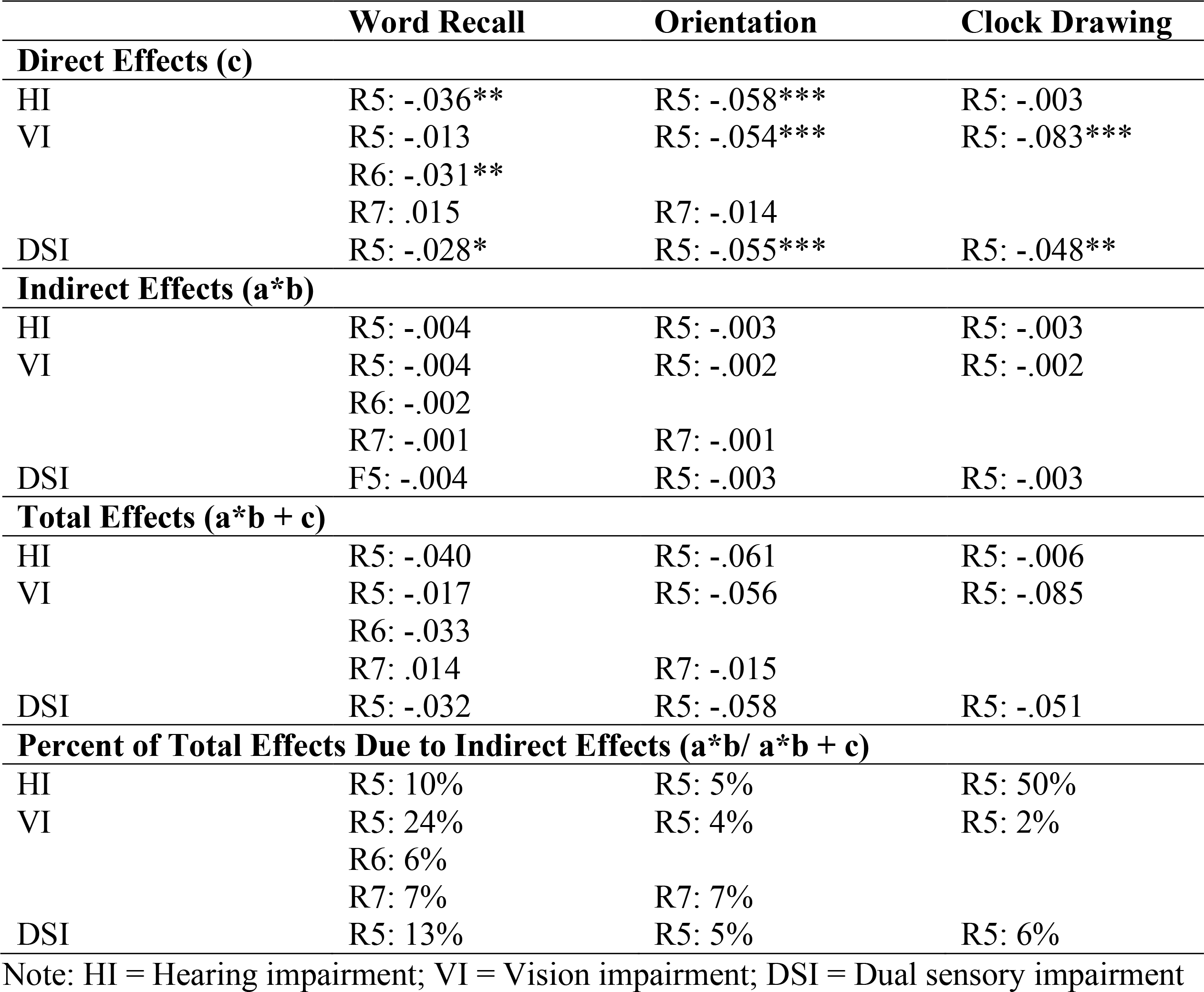
Decomposition of Effects, with Direct, Indirect, and Total Effects of Sensory Impairment Predicting Cognitive Functioning through Social Isolation.

